# Predicting 24-Month MCI-to-Alzheimer’s Conversion Using Routine Clinical Assessments Without Neuroimaging or Genetic Testing

**DOI:** 10.64898/2026.06.21.26356189

**Authors:** Sophie Choe

## Abstract

**INTRODUCTION:** Early identification of individuals with mild cognitive impairment (MCI) at high risk of conversion to Alzheimer’s disease (AD) is essential for timely intervention. We evaluated whether routinely obtainable clinical assessments can accurately predict 24-month MCI-to-AD conversion.

**METHODS:** Data from 2,430 participants with MCI in the Alzheimer’s Disease Neuroimaging Initiative were analyzed. XGBoost, Random Forest, and Logistic Regression models were evaluated. SHAP-based feature selection and feature ablation analyses assessed the incremental value of APOE ε4 genotype.

**RESULTS:** A six-feature model incorporating age, sex, education, RAVLT Immediate Recall, MMSE, and EcogSPTotal achieved an AUC of 0.922 (95% CI, 0.911–0.933). APOE ε4 provided negligible additional predictive value once cognitive measures were included. The XGBoost model outperformed Clinical Dementia Rating Sum of Boxes classification.

**DISCUSSION:** Routine cognitive assessments accurately predict 24-month MCI-to-AD progression without biomarkers, neuroimaging, or genetic testing, offering a practical, low-cost tool for clinical risk stratification.

## 1. Background

Mild cognitive impairment (MCI) affects approximately 15–20% of adults aged 65 years and older and represents the prodromal stage of Alzheimer’s disease (AD) for many individuals who eventually develop dementia [1–4]. However, progression is heterogeneous: while some individuals with MCI convert to AD within a few years, many remain cognitively stable or even revert to normal cognition [5,6]. Consequently, the primary clinical challenge is not identifying MCI itself, but determining which patients are at sufficiently high risk of near-term conversion to warrant closer monitoring, specialist referral, and advanced diagnostic evaluation.

The need for effective risk stratification has become increasingly important with the emergence of disease-modifying therapies for early symptomatic AD. Anti-amyloid monoclonal antibodies, including lecanemab and donanemab, have demonstrated the ability to slow cognitive decline in selected patients [7–9]. Early identification of individuals most likely to progress may therefore facilitate timely treatment planning, enrollment in clinical trials, and interventions targeting potentially modifiable risk factors.

Current biological models of AD increasingly emphasize biomarkers of amyloid pathology, tau pathology, and neurodegeneration [10,11]. These processes can be assessed using cerebrospinal fluid (CSF) assays [12,13], positron emission tomography (PET) [14], magnetic resonance imaging (MRI) [15], and emerging blood-based biomarkers [16]. Although these approaches have improved diagnostic precision, they often require specialized expertise, substantial cost, or infrastructure that may not be readily available in routine clinical practice. At the same time, healthcare systems face increasing demand for dementia evaluation, persistent underdiagnosis of MCI, and shortages of dementia specialists [5]. As a result, it is neither practical nor economically feasible to perform advanced diagnostic testing for every patient presenting with cognitive concerns.

These challenges underscore the need for scalable, low-cost, and easily administered tools capable of identifying individuals at highest risk of progression using information obtainable during a routine clinical encounter. An effective first-line risk stratification tool could support decision-making at the point of care, enabling primary care providers and other non-specialists to prioritize patients for closer monitoring, specialist referral, and resource-intensive confirmatory testing.

Machine learning approaches have shown considerable promise for predicting conversion from MCI to AD [17–26], but many published models rely on neuroimaging, fluid biomarkers, genetic testing, or extensive neuropsychological batteries unavailable during a routine clinical encounter. This limits their practical implementation despite strong research performance.

In this study, we developed and validated machine learning models to predict progression from MCI to AD within 24 months using assessments routinely obtainable during an initial evaluation. Initial models were constructed using demographic, cognitive, functional, and genetic variables from the Alzheimer’s Disease Neuroimaging Initiative (ADNI) [27]. Using SHAP-based feature selection [28], we identified the minimal set of variables required to preserve predictive performance. We then quantified the incremental contribution of APOE ε4 genotype [29] across successive levels of clinical information through systematic feature ablation. Finally, we compared model performance with the Clinical Dementia Rating Sum of Boxes (CDR-SB), a widely used measure of clinical disease severity [30].

## 2. Methods

The aim of this study was to develop a low-cost, easily administered risk stratification tool suitable for use in routine primary care settings. We trained machine learning models using demographic, cognitive, and genetic features, applied SHAP-based feature importance for selection, conducted ablation analyses to assess APOE4’s contribution, and benchmarked performance against CDR-SB.

### 2.1 Study Design

The primary outcome was conversion from MCI to AD within a 24-month follow-up window. The analytical pipeline followed five sequential stages: (1) algorithm comparison on a full feature set; (2) SHAP-based feature ranking; (3) retraining on a minimal feature set and comparison against full-feature models; (4) ablation analysis with and without APOE4; and (5) benchmarking against CDR-SB as a clinical reference standard.

### 2.2 Data Source

Data were obtained from the ADNI database (adni.loni.usc.edu). Launched in 2003, ADNI has enrolled participants across four phases (ADNI-1, ADNI-GO, ADNI-2, ADNI-3) under institutional review board approval at all participating sites [27]. Data are de-identified and publicly available to qualified researchers. After applying inclusion/exclusion criteria, 2,430 MCI participants were retained: 547 (22.5%) converted to AD within 24 months (progressors) and 1,883 did not (non-progressors), yielding a 3.4:1 class ratio.

### 2.3 Inclusion/Exclusion Criteria

Participants were included if they had a baseline MCI diagnosis, complete demographic and neuropsychological assessments, and sufficient longitudinal follow-up to determine 24-month progression status. Participants with baseline AD or cognitively normal status were excluded, as were those missing follow-up data, cognitive assessments, or APOE4 genotyping.

### 2.4 Features and Preprocessing

The candidate feature set comprised nine variables: three demographic measures (age, sex, years of education), APOE ε4 carrier status, and five clinical assessments. The clinical assessments included the Mini-Mental State Examination (MMSE), a 5–10 minute global cognitive screening tool [31]; the Everyday Cognition Scale Study Partner Total Score (EcogSPTotal), an informant-rated measure of everyday functional ability [32]; the Ecog Memory Discrepancy score (EcogMem_discrepancy), which captures the difference between self- and informant-rated memory functioning and may reflect anosognosia; RAVLT Immediate Recall, a verbal learning measure assessing the total number of words recalled across five learning trials [33]; and RAVLT Forgetting score, which quantifies the rate of delayed recall loss relative to peak learning performance. All features were selected to reflect assessments feasible during routine clinical encounters without specialized equipment or neuroimaging.

Continuous variables were standardized to numeric format. Missing values were imputed using training-fold medians and applied to test folds to prevent data leakage. Sex was binary-encoded (Male = 1, Female = 0). All preprocessing steps were implemented within the cross-validation loop using training-fold statistics exclusively, ensuring no information from held-out folds contaminated model training or evaluation.

### 2.5 Machine Learning Models

Three supervised classifiers were evaluated: Logistic Regression (LR) with L2 regularization and balanced class weighting, Random Forest (RF) [34] with 500 trees (max depth 4, minimum leaf size 5), and XGBoost [35] with 500 boosting rounds (max depth 3, learning rate 0.03). Hyperparameters were fixed a priori across all experiments to ensure fair comparison.

All models were evaluated using stratified 5-fold cross-validation with out-of-fold (OOF) prediction aggregation, ensuring every observation was evaluated exactly once by a model that had not seen it during training. Preprocessing used training-fold statistics only, preventing data leakage. Performance metrics — AUC, sensitivity, specificity, accuracy, and Brier score [36] — were computed on aggregated OOF predictions. Decision thresholds were optimized post-hoc using the Youden Index [37]. The Brier Skill Score (BSS) [38] quantified improvement over a naïve baseline. Class imbalance was addressed through balanced class weighting in LR and RF, and proportional scale_pos_weight in XGBoost.

### 2.6 Model Interpretability

To quantify feature contributions to model predictions, SHAP (SHapley Additive exPlanations) values were computed using TreeExplainer for the best-performing full model (XGBoost) across all out-of-fold (OOF) predictions. SHAP values decompose each individual prediction into additive feature contributions relative to the model’s baseline prediction, enabling patient-level interpretation while accounting for feature interactions.

Mean absolute SHAP values were used to rank features according to their average influence on predicted risk. Features with the highest importance were selected to construct the minimal feature set, thereby grounding dimensionality reduction in empirical predictive contribution rather than clinical assumption. SHAP beeswarm plots were generated to visualize the distribution and magnitude of feature effects across patients, and SHAP dependence plots were created for the top-ranked features to characterize the directionality of their associations with predicted risk.

### 2.7 Feature Ablation Design

The central analytic strategy was progressive feature ablation: model performance was evaluated as features were added incrementally in descending order of SHAP importance, mirroring the sequence in which clinicians typically obtain information. Four ablation levels were defined, and a separate XGBoost classifier was trained and evaluated at each level, with and without APOE4.

### 2.8 Clinical Baseline Comparator

CDR-SB was evaluated as a single-variable clinical reference. The Youden-optimal threshold (CDR-SB ≥ 2.0) was identified using the same ROC-based procedure applied to ML models. AUC, sensitivity, specificity, accuracy, and Brier score were computed on the full sample for direct comparison.

### 2.9 Statistical Analysis

AUC differences between ablation models were assessed using DeLong’s method [39] for correlated ROC curves, with differences below 0.01 pre-specified as the threshold for practical equivalence. Analyses were conducted in Python 3.12 using pandas, NumPy, scikit-learn v1.4, XGBoost v2.0, and SHAP v0.44. This study is reported in accordance with the TRIPOD statement [40].

## 3. Results

### 3.1 Full Model Comparison

All three algorithms demonstrated strong discriminative performance on the full nine-feature set (Table 1). Random Forest achieved the highest AUC, while XGBoost demonstrated the most favorable balance across discrimination, sensitivity, and calibration — including the highest sensitivity and Brier Skill Score. Logistic Regression performed comparably across all metrics.

**Table 1.**
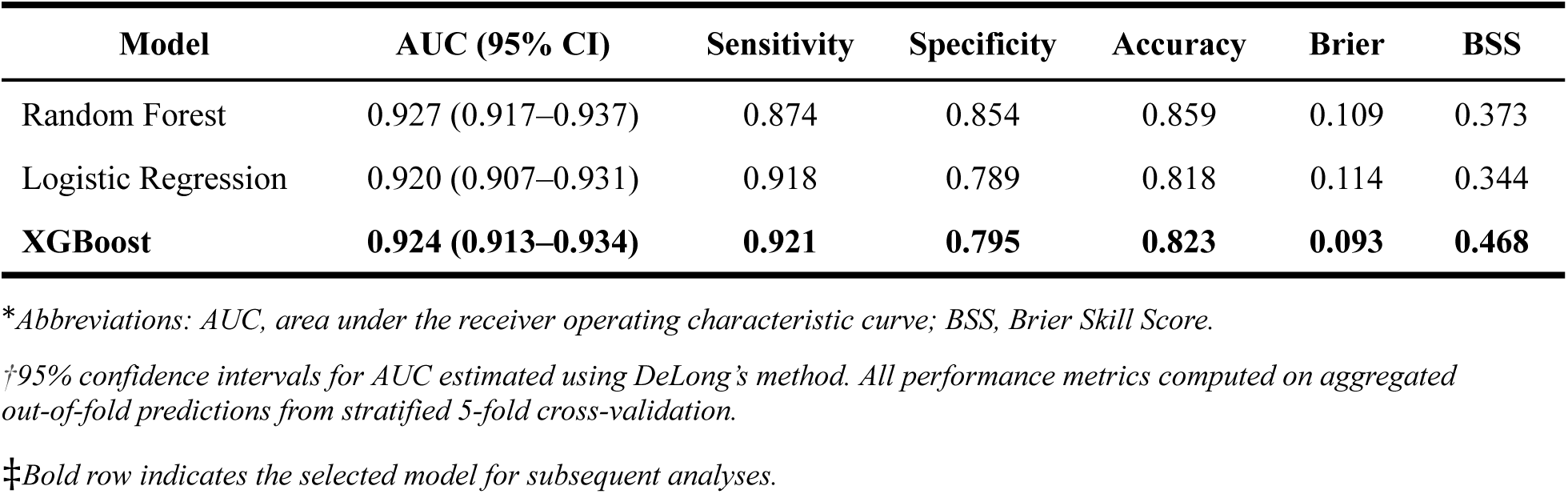
Model Performance on the Full Feature Set.

XGBoost was selected for subsequent SHAP analysis based on its superior calibration, highest BSS, and greatest sensitivity, reflecting the clinical priority of minimizing missed conversions.

### 3.1 SHAP Feature Importance

SHAP analysis of the full XGBoost model identified three features as dominant contributors to predicted conversion risk. RAVLT Immediate Recall (mean |SHAP| = 1.26), MMSE (1.16), and EcogSPTotal (1.12) exhibited substantially greater importance than all other variables, with mean absolute SHAP values approximately threefold higher than the remaining features. Among the secondary predictors, APOE4 ranked fourth (0.40), followed by EcogMem Discrepancy (0.30), RAVLT Forgetting (0.25), Age (0.23), Education (0.19), and Sex (0.09).

The SHAP beeswarm plot (Figure 1) demonstrated consistent directional effects for the three most influential features.

**Figure 1.**
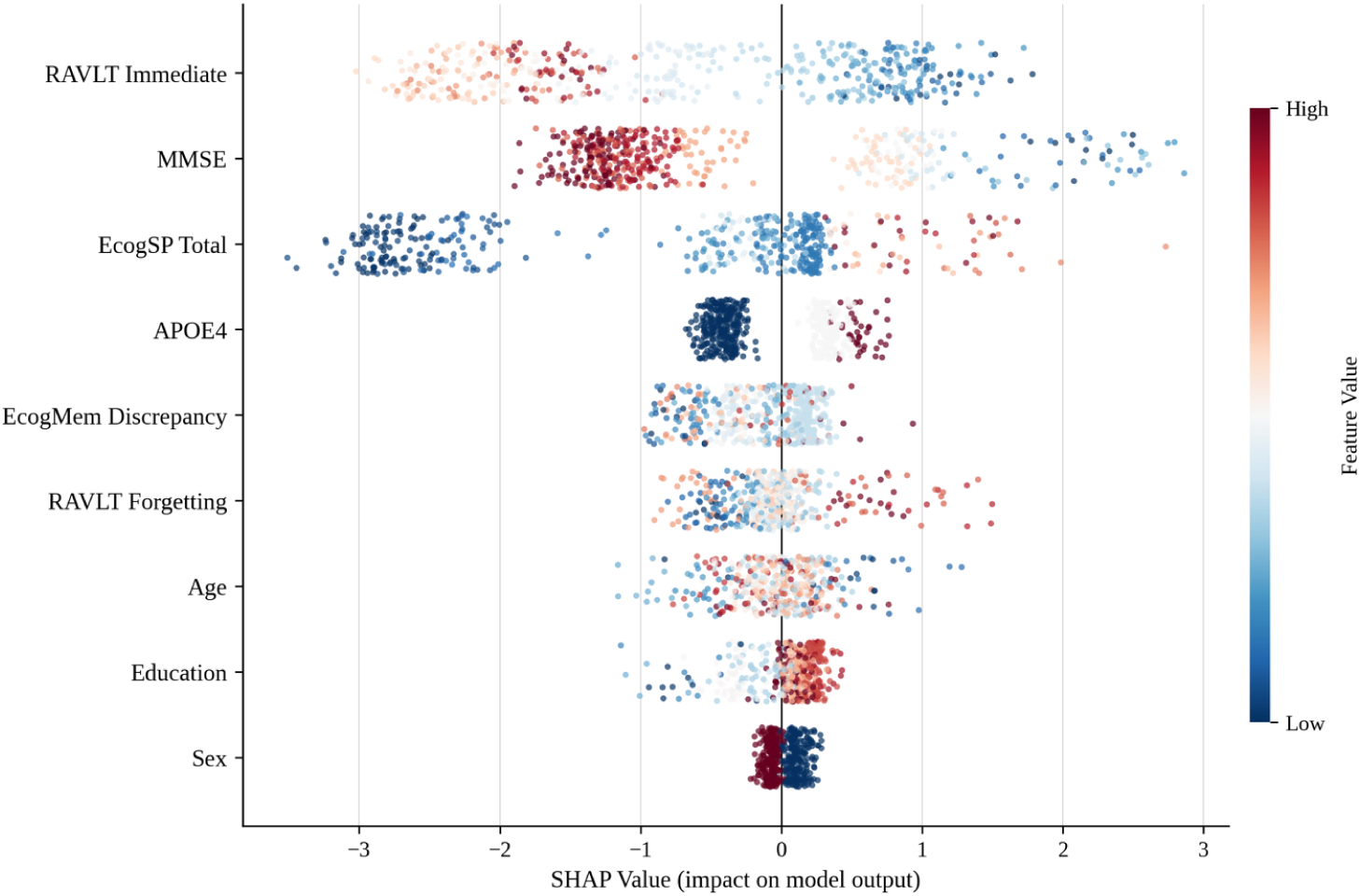
SHAP Beeswarm Plot of Feature Contributions to Individual Predictions.

Higher RAVLT Immediate Recall and MMSE scores were associated with lower predicted risk of progression, whereas poorer performance shifted predictions toward higher risk. Similarly, higher EcogSPTotal scores, reflecting greater informant-reported functional decline, increased predicted conversion risk. Although APOE4 carrier status separated individuals into higher- and lower-risk groups, its contribution was substantially smaller than those of the leading cognitive and functional measures. The remaining features showed limited dispersion around zero, indicating relatively modest and inconsistent effects on individual predictions.

The marked decline in SHAP importance after the top three features supported the development of a parsimonious model emphasizing variables that are routinely collected in clinical practice. Accordingly, Age, Sex, Years of Education, RAVLT Immediate Recall, MMSE, and EcogSPTotal were selected for the six-feature minimal model evaluated in Stage 3. This feature set balanced predictive performance with clinical feasibility by relying exclusively on demographic, cognitive, and functional assessments that require no specialized testing or equipment.

### 3.2 Minimal Model Comparison

Retraining all three algorithms on the six-feature minimal set (age, sex, education, RAVLT Immediate Recall, MMSE, and EcogSPTotal) resulted in virtually no loss of predictive performance. For XGBoost, AUC decreased by only 0.002, from 0.924 (95% CI: 0.913–0.934) in the full model to 0.922 (95% CI: 0.911–0.933) in the minimal model. Calibration was preserved, and both Brier Skill Score and specificity improved slightly. Random Forest and Logistic Regression showed similarly stable performance. These results demonstrate that a parsimonious model based on six routinely obtainable features captures nearly all of the predictive information contained in the full nine-feature model.

**Table 2.** Model Performance on the Minimal Feature Set Compared with CDR-SB.

| Model | AUC (95% CI) | Sensitivity | Specificity | Accuracy | Brier | BSS | Features |
| --- | --- | --- | --- | --- | --- | --- | --- |
| Random Forest | 0.921 (0.910–0.932) | 0.912 | 0.802 | 0.827 | 0.114 | 0.347 | 6 |
| Logistic Regression | 0.915 (0.903–0.927) | 0.872 | 0.828 | 0.838 | 0.117 | 0.328 | 6 |
| **XGBoost** | **0.922 (0.911–0.933)** | **0.878** | **0.836** | **0.845** | **0.093** | **0.470** | **6** |
| CDR-SB ($\geq 2.0$) | 0.912 | 0.830 | 0.830 | 0.830 | 0.117 | 0.330 | 1 |

\*Abbreviations: AUC, area under the receiver operating characteristic curve; BSS, Brier Skill Score; CDR-SB, Clinical Dementia Rating Sum of Boxes.
†95% confidence intervals for AUC estimated using DeLong's method for ML models; CI not reported for CDR-SB as it was evaluated as a fixed clinical threshold on the full sample rather than via cross-validation.
‡All ML model metrics computed on aggregated out-of-fold predictions from stratified 5-fold cross-validation. Bold row indicates the best-performing minimal ML model.

The reduction from nine to six features produced minimal impact on predictive accuracy across all three algorithms, confirming that the three excluded variables — APOE4, EcogMem Discrepancy, RAVLT Forgetting— contributed negligible incremental value beyond the retained feature set. The six-feature XGBoost model was consequently selected for subsequent ablation analyses.

### 3.3 Feature Ablation Analysis

Feature ablation revealed rapidly diminishing marginal returns as cognitive variables were added to the demographic baseline (Table 3). APOE4 provided substantial predictive value when only demographic information was available, increasing AUC from 0.524 to 0.651 (+0.127). However, the addition of RAVLT Immediate Recall produced the largest improvement in model performance, increasing AUC to 0.855 (+0.331) without APOE4. Thereafter, the incremental contribution of APOE4 declined sharply from 0.1269 to 0.011 following addition of RAVLT Immediate Recall and to 0.008 following addition of MMSE.

**Table 3.** Incremental Contribution of APOE4 Genotype Across Sequential Feature Sets.

| Features<br>Demographics + | AUC | AUC<br>(+APOE4) | APOE4<br>Contribution |
| --- | --- | --- | --- |
| None | 0.5242 | 0.6511 | +0.1269 |
| RAVLT | 0.8554 | 0.8668 | +0.0114 |
| RAVLT + MMSE | 0.8923 | 0.9004 | +0.0081 |
| RAVLT + MMSE + EcogSPTotal | 0.9123 | 0.9113 | -0.0010 |

\*Abbreviations: AUC, area under the receiver operating characteristic curve; APOE4, apolipoprotein E ε4; RAVLT, Rey Auditory Verbal Learning Test (immediate recall); MMSE, Mini-Mental State Examination; EcogSPTotal, Everyday Cognition Study Partner Total Score.
†Demographics include age, sex, and years of education.
‡APOE4 contribution is calculated as AUC with APOE4 minus AUC without APOE4 at each feature level.

With the complete minimal feature set, model performance reached an AUC of 0.912 without APOE4. Inclusion of APOE4 yielded no further improvement (AUC = 0.911), indicating that genetic information was fully redundant once routine cognitive and functional assessments were available.

These findings highlight two important observations. First, RAVLT Immediate Recall was the single most influential predictor, accounting for the majority of performance gains beyond demographics. Second, the predictive value of APOE4 was highly context-dependent: while informative in the absence of cognitive data, its contribution became negligible once standard neuropsychological assessments were incorporated, suggesting limited utility for short-term progression risk stratification after routine clinical evaluation.

### 3.4 CDR-SB Comparison

To benchmark the minimal model against established clinical practice, CDR-SB was evaluated as a single-variable comparator using a Youden-optimal threshold of ≥ 2.0. CDR-SB achieved an AUC of 0.912, sensitivity of 0.830, specificity of 0.830, and Brier score of 0.117. The six-feature XGBoost minimal model exceeded CDR-SB performance across all discrimination and calibration metrics (AUC 0.922 vs. 0.912; Brier score 0.093 vs. 0.117; BSS 0.470 vs. 0.330), while additionally offering a structured, reproducible probabilistic output rather than a threshold-based binary classification dependent on a clinician-administered interview.

## 4. Discussion

This study developed a machine learning model for predicting 24-month MCI-to-AD conversion using a systematically reduced set of routinely obtainable clinical measures. Three principal findings emerged: high predictive performance (AUC > 0.92) is achievable with as few as six features; verbal memory and global cognitive status account for the dominant share of predictive signal; and APOE4 genotype, while informative in the absence of cognitive data, becomes fully redundant once a brief neuropsychological assessment is available.

### 4.1 Model Performance and Algorithm Selection

All three algorithms achieved strong and broadly comparable discrimination on the full feature set (AUC 0.920–0.927), consistent with prior literature showing that performance differences between well-tuned algorithms on structured tabular clinical data are typically small [41]. XGBoost was selected for its superior calibration (Brier score = 0.093, BSS = 0.468) and highest sensitivity (0.921), reflecting the clinical priority of minimizing missed conversions. Performance was stable across five stratified folds (CV AUC = 0.9245 ± 0.0125), and the consistency between mean CV AUC and OOF AUC (0.9244) confirms that reported metrics are unbiased estimates of out-of-sample performance.

### 4.2 Cognitive Predictors Dominate Conversion Risk

SHAP analysis identified RAVLT Immediate, MMSE, and EcogSPTotal as the three features most strongly driving predictions, with mean absolute SHAP values approximately three times larger than any remaining feature. The prominence of immediate verbal recall aligns with established evidence that episodic memory encoding is among the earliest and most sensitive markers of incipient AD pathology [42]. MMSE reflects the global cognitive decline characterizing MCI-to-AD progression, while EcogSPTotal captures functional consequences that self-report measures may underestimate due to anosognosia. Together, these measures mirror the dual-domain evaluation recommended by current MCI diagnostic frameworks [43], suggesting the model’s feature structure is both statistically optimal and conceptually aligned with clinical practice. Notably, RAVLT Forgetting contributed less than RAVLT Immediate, suggesting that encoding deficit rather than accelerated forgetting rate is the more discriminating signal at the MCI stage.

### 4.3 Feature Reduction Without Performance Cost

Reducing from nine to six features resulted in an AUC decline of just 0.002 for XGBoost, with BSS marginally improving from 0.468 to 0.470, suggesting the three excluded features introduced noise rather than signal. The near-identical performance of all three algorithms on the minimal feature set confirms that the six retained features capture the substantive predictive structure of the data.

The comparison with CDR-SB is particularly instructive. Despite being the gold standard staging measure in AD clinical trials, CDR-SB requires a structured clinician-administered interview taking 20–30 minutes. The minimal model exceeded CDR-SB on AUC (0.922 vs. 0.912), Brier score (0.093 vs. 0.117), and BSS (0.470 vs. 0.330), while relying on faster, less training-intensive measures.

### 4.4 The Diminishing Value of APOE4

The ablation analysis yielded one of the study’s most clinically relevant findings: the predictive value of APOE ε4 was highly dependent on the amount of clinical information available. When only demographic variables were included, APOE ε4 status improved discrimination substantially (ΔAUC = +0.127). This contribution declined to 0.011 following the addition of RAVLT Immediate Recall and disappeared entirely (ΔAUC = −0.001) in the complete six-feature model. These findings suggest that APOE ε4 testing may be most useful in settings where cognitive assessment is unavailable, such as population-based screening or initial triage in resource-limited environments. Once a brief neuropsychological evaluation has been completed, genetic testing appears to provide little additional value for short-term conversion prediction as seen in Figure 2.

**Figure 2.**
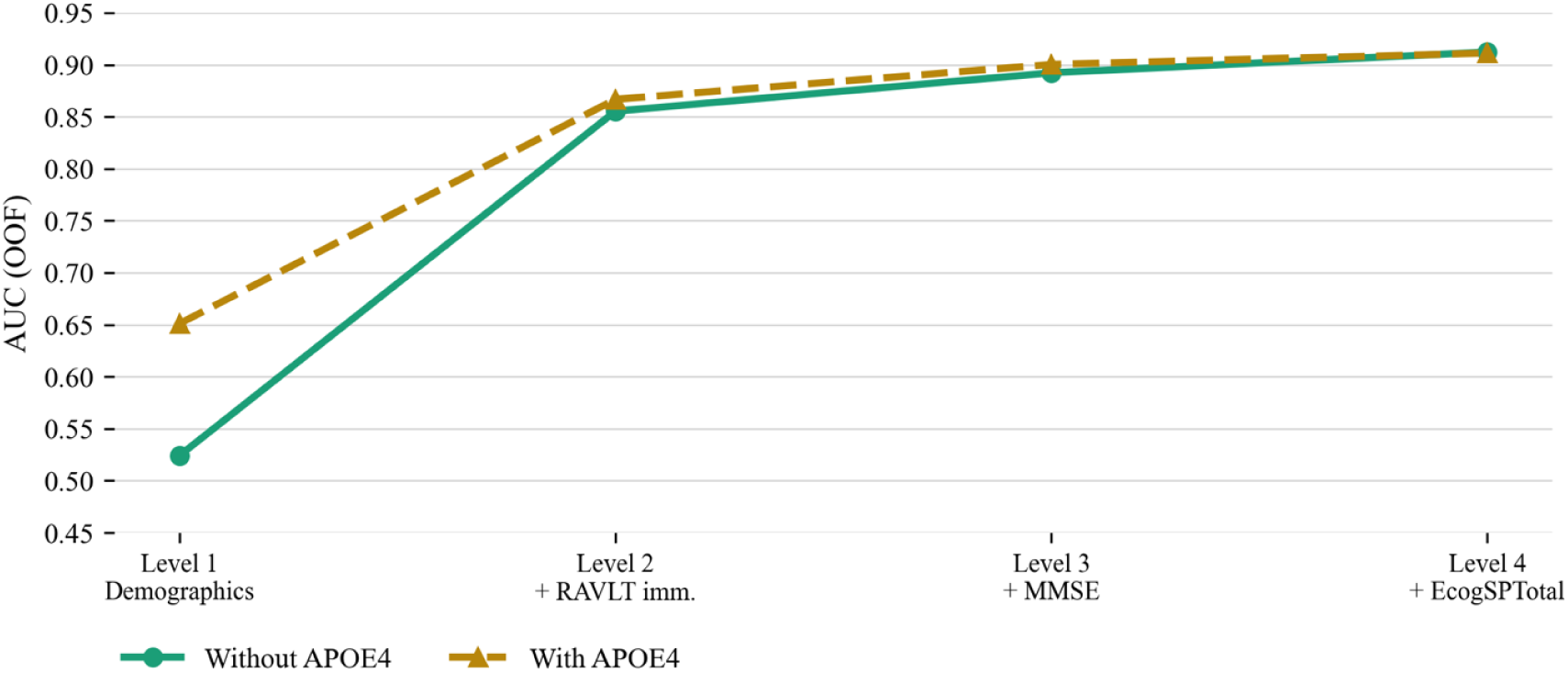
Feature Ablation Analysis: Marginal Contribution of APOE4 Across Sequential Levels of Clinical Information.

The ability to achieve comparable performance using routine assessments alone has important clinical implications, given that APOE ε4 testing introduces practical, ethical, and financial considerations including genetic counseling needs, patient anxiety, and insurance discrimination concerns.

### 4.5 Comparison with Published Models

Previous machine learning models predicting MCI-to-AD progression have frequently relied on structural MRI, PET imaging, CSF biomarkers, genetic information, or multimodal combinations, reporting AUC values of approximately 0.74–0.93 in ADNI-based cohorts [17–26]. The six-feature model achieved comparable discrimination (AUC = 0.922; 95% CI: 0.911–0.933) using only demographic, cognitive, and functional assessments obtainable during a routine clinical visit.

The principal contribution is therefore not a higher AUC, but the demonstration that strong predictive performance is achievable without neuroimaging, fluid biomarkers, or genetic testing. Multimodal approaches may provide incremental gains in discrimination but require specialized infrastructure, higher costs, and expert interpretation — none of which are necessary here. Table 4 summarizes the practical tradeoffs associated with commonly used approaches, and highlights the accessibility of the features retained in the present model. From a clinical perspective, such a tool is particularly valuable as a first-line stratification instrument, identifying patients who warrant closer monitoring, specialist referral, or confirmatory testing.

**Table 4.** Cost, Equipment, and Availability of Candidate Assessments and Biomarkers for MCI-to-AD Risk Stratification.

| Test | Cost | Equipment Required | Typical Availability |
| --- | --- | --- | --- |
| MMSE | Very low | None | Very common |
| ECog | Very low | None | Common |
| RAVLT | Low | None | Common in memory clinics |
| APOE4 genotyping | Moderate | Genetic testing | Variable |
| Plasma p-tau217 | Moderate | Specialized assay | Emerging |
| CSF Aβ42/tau | Higher | Lumbar puncture | Specialist centers |
| Amyloid PET | High | PET scanner | Limited |
| Tau PET | Very high | PET scanner | Limited |
| MRI volumetrics | Moderate–high | MRI + processing | Variable |

*\*Abbreviations: MMSE, Mini-Mental State Examination; ECog, Everyday Cognition; RAVLT, Rey Auditory Verbal Learning Test; APOE4, apolipoprotein E ε4; CSF, cerebrospinal fluid; Aβ42, amyloid-beta 42; PET, positron emission tomography; MRI, magnetic resonance imaging.*
*†Cost and availability estimates reflect general clinical practice in the United States and may vary by region and healthcare setting. Assessments categorized based on published guidelines and consensus statements [5, 16, 30, 42].*

### 4.6 Limitations

Several limitations warrant consideration. ADNI participants are predominantly White, highly educated, and academically recruited, limiting generalizability to diverse primary care populations. Performance was evaluated within a single dataset, and independent external validation is necessary to establish transportability. The 24-month prediction horizon was fixed; performance across shorter or longer windows remains unexplored. APOE4 redundancy findings are specific to XGBoost and may not generalize across modeling approaches. Finally, EcogSPTotal relies on informant reports and may be influenced by informant characteristics or anosognosia.

### 4.7 Future Directions

External validation across diverse populations and real-world primary care settings is the immediate priority, as ADNI’s cohort limits generalizability to community-based clinical environments. Future studies should evaluate nonspecialist administration in settings where biomarker and neuroimaging infrastructure is unavailable or cost-prohibitive — the deployment context for which this tool is most directly intended. Longitudinal applications represent a natural extension: repeated administration across clinical visits could enable trajectory-based monitoring of conversion risk over time, transforming the tool from a one-time screening instrument into a dynamic monitoring companion relevant to the era of disease-modifying therapies. The model may additionally serve as a first-line stratification layer within staged diagnostic frameworks, identifying individuals most likely to benefit from referral for confirmatory plasma biomarker, CSF, or amyloid PET evaluation — supporting more efficient resource allocation and clinical trial enrollment.

### 4.8 Conclusion

This study demonstrates that progression from MCI to AD within 24 months can be predicted with high accuracy using six features routinely obtainable during a single clinical encounter, without neuroimaging or genetic testing. These findings support the development of accessible, low-cost prediction tools deployable across diverse healthcare settings where biomarker-intensive approaches are impractical or unavailable.

## Data Availability

Data used in this study were obtained from the Alzheimer's Disease Neuroimaging Initiative (ADNI) at https://adni.loni.usc.edu/. The code supporting the analyses reported in this study, including data preprocessing, model training, evaluation, SHAP analyses, and feature ablation experiments, is publicly available at https://github.com/Clinovia/mci-ad-prediction.

https://github.com/Clinovia/mci-ad-prediction

## Author Approval

All the authors have seen and approved the manuscript.

## Ethics Statement

This study used de-identified, publicly available data obtained from the Alzheimer’s Disease Neuroimaging Initiative (ADNI). The original ADNI study received approval from institutional review boards at participating sites, and all participants provided written informed consent. The present secondary analysis of de-identified data did not require additional institutional review board approval.

